# Cerebral Small Vessel Disease and Risk of Cardiovascular Events and All-Cause Mortality: The Framingham Heart Study

**DOI:** 10.64898/2026.09.01.26361997

**Authors:** Riya Manchanda, Adlin Pinheiro, Hugo J. Aparicio, Vasileios Lioutas, Alexa S. Beiser, Joseph Sisto, Oluchi Ekenze, Emelia J. Benjamin, Charles DeCarli, Sudha Seshadri, Jose R. Romero

**Affiliations:** Department of Neurology, Boston University Chobanian & Avedisian School of Medicine; Department of Biostatistics, Boston University School of Public Health; NHLBI’s Framingham Heart Study; Department of Neurology, Beth Israel Deaconess Medical Center; Section of Cardiovascular Medicine, Boston Medical Center, Boston University Chobanian & Avedisian School of Medicine, Department of Epidemiology, Boston University School of Public Health; Department of Neurology, University of California at Davis; The Glenn Biggs Institute for Alzheimer’s and Neurodegenerative Diseases, University of Texas Health Sciences Center

## Abstract

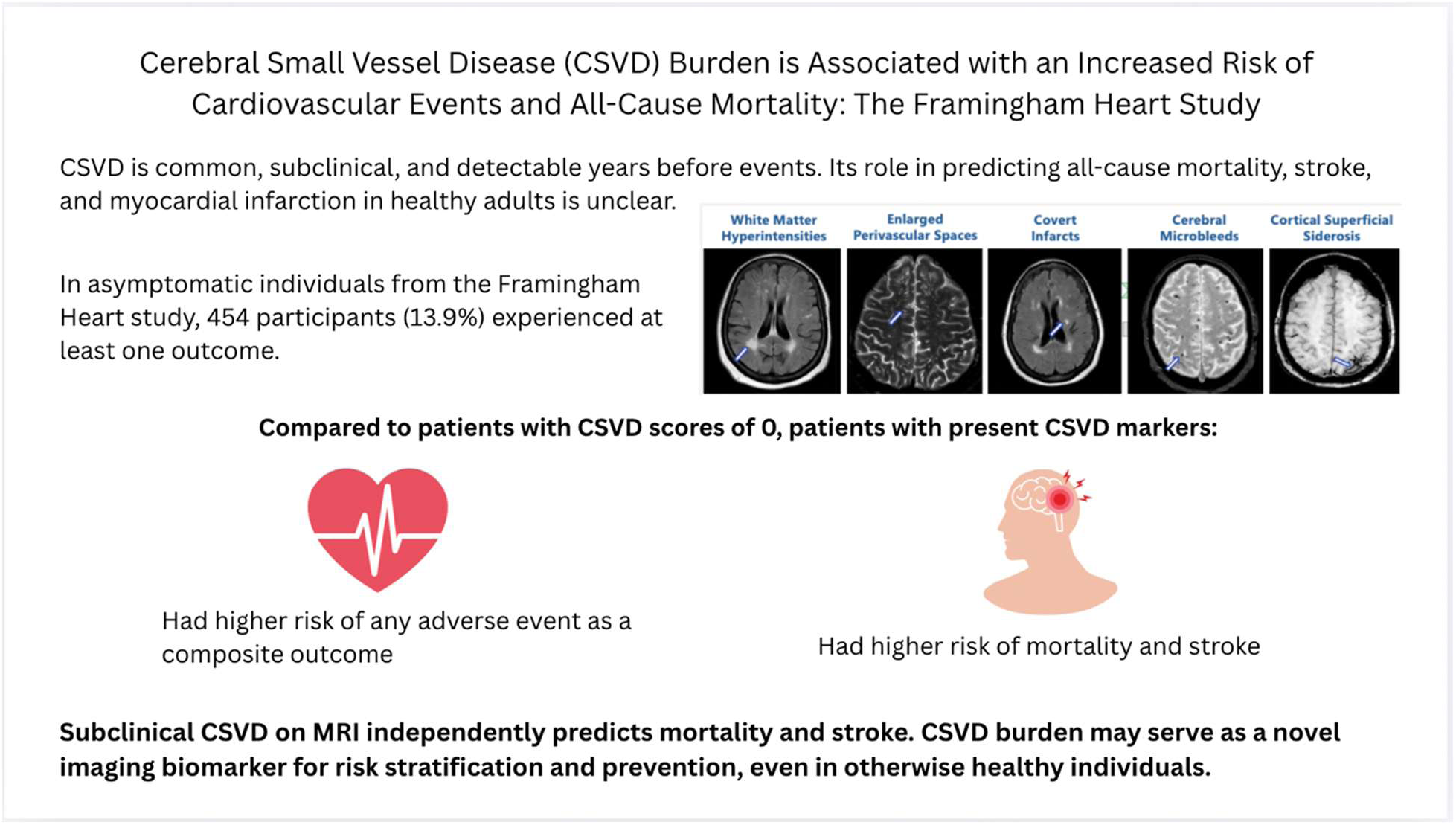

**BACKGROUND:** The effect of neurovascular health on cardiovascular outcomes is not well-known. Cerebral small vessel disease (CSVD) signals subclinical vascular brain injury and its quantification may serve as a tool to stratify risk in healthy individuals. Studying the relations of CSVD to cardiovascular events and mortality could aid in targeted preventive efforts.

**METHODS:** We included Framingham Heart Study participants free of diagnosed stroke and dementia. CSVD was assessed on brain MRI—considering covert infarcts, cerebral microbleeds, cortical superficial siderosis, high perivascular space burden, and extensive white matter hyperintensities—with 1 point assigned for each marker, to categorize CSVD scores as 0, 1, or 2+. We used a composite outcome and three components: stroke, myocardial infarction, and mortality. Cox regression analysis was used with models adjusted for age, sex, cohort, MRI-exam cycle time interval, and vascular risk factors.

**RESULTS:** In our study, 31% of our 3263 participants (mean age 58.0 years [SD: 13.4], 54% women) had a CSVD score greater than 0, and those with higher scores were generally older and had higher prevalence of vascular risk factors. Over a median follow-up of 8.4 years, 454 participants (13.9%) experienced at least one outcome, with mortality being the most common. Increasing CSVD burden was associated with higher incidence rates of the composite outcome. Compared with participants with CSVD score 0, those with scores of 1 and 2 had significantly higher risk of any event (HR 1.48, 95% CI 1.19–1.85; HR 1.52, 95% CI 1.17–1.97 respectively). CSVD burden showed a dose-response relationship with all-cause mortality and was associated with increased stroke risk, while no significant association with MI was observed.

**CONCLUSION:** Higher subclinical CSVD burden is independently associated with increased risk of adverse cardiovascular events and mortality in community-dwelling individuals. CSVD detected on brain MRI may serve as a valuable imaging biomarker for risk stratification and early prevention of life-threatening outcomes.

Indexing Terms: Stroke, Mortality, Myocardial Infarction, Cerebral Small Vessel Disease, Vascular Disease, Brain MRI

## Introduction

Cerebral small vessel disease (CSVD) is a highly prevalent neurovascular pathology among middle-aged and older patients. CSVD is heterogeneous, composed of several clinical, pathological, and neuroimaging features, and results from diseased cerebral vasculature, including arterioles, capillaries, and venules.^1^ CSVD can be readily assessed on brain magnetic resonance imaging (MRI) using markers such as white matter hyperintensities, cerebral microbleeds, MRI-visible perivascular spaces, cortical microinfarcts, and brain atrophy.^2^ In addition, CSVD contributes heavily to vascular cognitive impairment^3^ and dementia,^4^ and is associated with functional and cognitive decline, urinary disturbances,^5^ gait disorders,^6^ and stroke. CSVD is responsible for 25% of ischemic strokes and most hemorrhagic strokes.^7,8^ Its presence is also associated with worse prognosis after stroke, including impaired recovery^9^ and increased recurrence.^10^

Advanced age is strongly related to CSVD,^7,11^ with prevalence rising from 5% in people 50 years of age to almost 100% in people above 90 years.^12^ Thus, as the population ages, the impact of CSVD is expected to increase. Additionally, several modifiable risk factors such as hypertension, smoking, hyperlipidemia, and diabetes are strongly related to CSVD.^13,14^ Since CSVD predates clinical neurological events by years to decades, it is thus a preventive treatment target and can be used to stratify intervention as well.

Although increased risk for stroke in relation to CSVD is well established, further study is needed to characterize CSVD burden with overall cardiovascular and mortality risk. Prior studies have examined individuals with previously existing disease in smaller populations.^15^ Limited data are available in healthy individuals dwelling in the community. In addition, a composite score of CSVD encompasses multiple markers that may reflect the cumulative exposure to vascular risk factors. Rather than using assessments of individual risk factors, a composite score may more accurately reflection CSVD burden in individuals. Studying this relationship may elucidate potential cerebrovascular contributions to overall health. It may also support using imaging-detected CSVD burden, even in subclinical stages of disease, to identify individuals at higher risk and guide targeted treatment. Therefore, we studied the association between a multi-marker CSVD score capturing overall burden, as measured by a composite score of neuroimaging features, and incidence of adverse outcomes in healthy community-dwelling individuals.

## Methods

We followed the STROBE guidelines for reporting of our study.

### Sample (description, inclusion, exclusion criteria)

Participants from the Original, Offspring, Third Generation, Omni Cohort 1, and New Offspring Spouse (NOS) cohorts of the Framingham Heart Study (FHS) were eligible for inclusion. The Original cohort (Gen 1, N = 5,209) was first enrolled in 1948 and underwent biennial examinations through 2014, totaling 32 exam cycles. The Offspring cohort (Gen 2, N = 5,124), composed of the children of the Original cohort participants and their spouses, was launched in 1971 and has been evaluated approximately every 4–7 years; it has just completed its 10th cycle. The Omni Generation 1 cohort (N = 507), established in 1994 to better represent the area’s increasing diversity, includes primarily individuals of African American, Hispanic, Asian, Indian, Pacific Islander, and Native American descent. Examinations for this group have been held every 4-8 years and has completed their 5th cycle in 2022. The NOS cohort, which includes spouses of the Offspring generation, and the Generation 3 cohort (Gen 3, N = 4,095), consisting of the children of the Offspring cohort, were introduced in 2002-2003 and participated in exams every 5–8 years. They are now in their 4th exam cycles.

FHS participants were eligible for the present study if they attended at least one clinic exam at which MRIs were obtained and had ratings present for all CSVD markers. Of the 9,147 eligible participants, 3,710 had at least one MRI that was rated for CSVD markers. If multiple MRIs were available, the earliest was selected for analysis. Participants were excluded if they had prevalent dementia, history of stroke, myocardial infarction (MI), or other neurological conditions (e.g. head injury, tumors) at the time of MRI, or were missing baseline covariate data or follow-up data for either mortality, MI, or stroke. Brain MRIs were obtained between June 1999 and August 2015. The follow-up period spans from the time of brain MRI until December 31, 2020. The sample selection process is illustrated in Figure 1.

**Figure 1.**
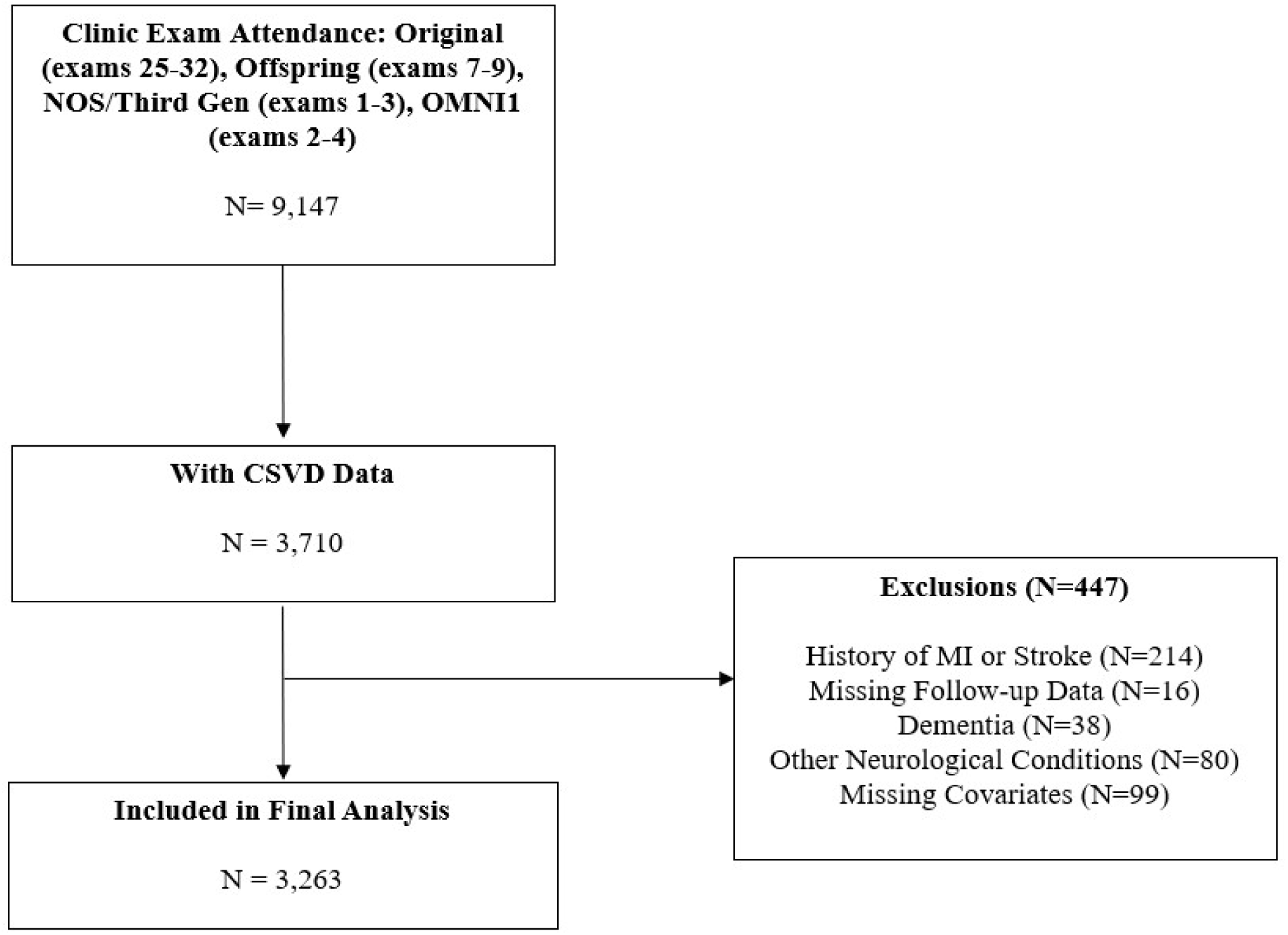
Sample selection flowchart.

The Institutional Review Board of Boston University Medical Campus approved the study protocol and informed consent was obtained from all subjects.

### Brain MRI

Brain MRI measures and acquisition methods have been previously described in detail.^16,17^ Participants underwent MRI on either a 1T (Siemens Magnetom; 1999-2004) or 1.5T scanner (GE Signa or Siemens Avanto/Symphony; 2001-2025). MRI protocols included sequences allowing for ratings of CSVD markers.

### Cerebral small vessel disease (CSVD)

CSVD burden was defined using a composite score of five neuroimaging features seen on MRI scans, assigning one point to each present feature: extensive white matter hyperintensities (WMH), covert brain infarcts (CBI), MRI-visible perivascular spaces (PVS), cerebral microbleeds (CMB), and cortical superficial siderosis (cSS). Each MRI feature was defined using the consensus criteria by the Standards for Reporting Vascular Changes on Neuroimaging Criteria (STRIVE consortium).^7^

CBI were defined by their size and appearance on T2-weighted sequences based on previously used methods.^18^ Lesions larger than 3 mm were considered infarcts. Lesions were also required to have cerebrospinal fluid density on subtraction images and to be separate from the circle of Willis vessels. Inter and intra-rater reliability measurements were evaluated by three different raters, with agreement values between 0.73 and 0.90.^18^

T2 weighted sequences were used to determine PVS burden in the basal ganglia and centrum semiovale. Details have been previously reported.^19^ Briefly, PVS signal intensity was similar to that of cerebrospinal fluid, with a pattern of penetrating vessels (linear when parallel to the penetrating vessel or round/ovoid when perpendicular to the penetrating vessel), and a diameter less than 3 mm. PVS burden was categorized into grades I-IV in each region based on counts: grade I (1-10), grade II (11-20), grade III (20-40) and grade IV (> 40). Grades III-IV in either the basal ganglia or centrum semiovale were considered high burden for this study. The inter-rater and intra-rater values were high for PVS, ranging from good to excellent in the basal ganglia and centrum semiovale, with ICC values of above 0.75.^20^

WMH volume was measured with the appropriate sequences (fluid attenuated inversion recovery or dual spin-echo) and volume was determined according to previously published methods.^16^ Extensive WMH volume was defined greater than one standard deviation above an age-group specific mean, expressed as a percentage of total brain volume. More details are reported elsewhere.^21^ For WMH volume, reliability measurements all yielded values above 0.90.^22^

CMB presence was analyzed using published guidelines.^23^ CMB were defined as rounded or ovoid hypointense lesions on T2*-GRE–weighted sequence. The lesions measured 10 mm or less in diameter and were surrounded by brain parenchyma over at least half their circumference. CMB showed excellent reliability measures, with kappa values of 0.78 for inter and intra-rater reliability.^24^

cSS was rated as per methods described elsewhere.^25–27^ Briefly, cSS was defined as linear, gyriform areas of low signal intensity along the superficial layers of the cerebral cortex, without corresponding hyperintensity on T1-weighted or FLAIR images and not contiguous with any intracerebral hemorrhage. Assessments performed on cSS presence and severity showed sufficient kappa values for reliability.^25^

### Outcomes

The protocols for clinical event surveillance and assessment for FHS have been published previously.^28^ FHS participants are monitored regularly for the occurrence of vascular events and death. The surveillance process entails not only questions related to cardiovascular events during each routine follow-up FHS clinic visit, but also mailed health history update questionnaires, which include detailed sections about interim events and hospitalizations. If interim events were reported, all applicable medical records were collected and reviewed by an events adjudication committee. This consisted of a panel of three investigators who reviewed hospital and clinic data, office visit notes, and lab and pathology reports.

We included all types of strokes for this study. The diagnosis for stroke was determined by a panel of investigators that included at least two neurologists.^29^ Stroke ascertainment was based on review of all clinical records, imaging, and autopsy when available. Stroke was defined as acute-onset focal neurologic deficits of presumed vascular etiology lasting more than 24 hours. Transient ischemic attacks followed the same definition with symptoms lasting less than 24 hours and were included in the definition for stroke.

MI diagnosis was done by a panel of physicians as described previously. MI was defined when the participant had at least two of the following: (1) symptoms indicative of ischemia, such as chest discomfort, (2) changes in blood biomarkers of myocardial necrosis, and (3) serial changes in the electrocardiogram^30^.

Ongoing surveillance for mortality is conducted at examination cycles through examinations and health updates. Additionally, deaths occurring outside of these cycles may be reported by family members to Framingham Heart Study staff. Once a death event is captured, it is reviewed by a panel to ascertain the cause of death, considering clinic records, physician notes, interviews with surviving family members, and autopsy information^31^. We considered death of any cause for this study.

### Vascular Risk Factors

Current cigarette smoking was defined as self-reported use in the year prior to the examination. Systolic (SBP) and diastolic (DBP) blood pressures were each taken as averages of 2 measurements from the Framingham clinic physician. Hypertension status was evaluated using the JNC-7 classification (SBP ≥ 140 mmHg or DBP ≥90 mmHg or usage of antihypertensive medications).^32^ We defined diabetes as a random blood glucose ≥ 200 mg/dl for the Original cohort, fasting glucose ≥ 126 mg/dl (≥ 7 mmol/L) for the Offspring, Third Generation, NOS, and Omni 1 cohorts, or use of insulin or oral hypoglycemic medications (for all cohorts). Body mass index (BMI) was calculated by the ratio of the participants’ weight and squared height (kg/m^2^). Total cholesterol was measured on fasting specimens in the Offspring cohort, and random samples in the Original cohort. Medication use was assessed by self-report.

### Statistical Analysis

Baseline characteristics of study participants were reported as frequencies and percentages for categorical variables and means and standard deviations for continuous variables. In multivariable analyses, CSVD scores greater than 2 were grouped together due to small sample sizes, resulting in score categories of 0, 1, and 2+. Further sensitivity analyses were also conducted using groups of 0, 1, 2, and 3+ and are reported in Supplemental Material Table 3.

**Table 1.**
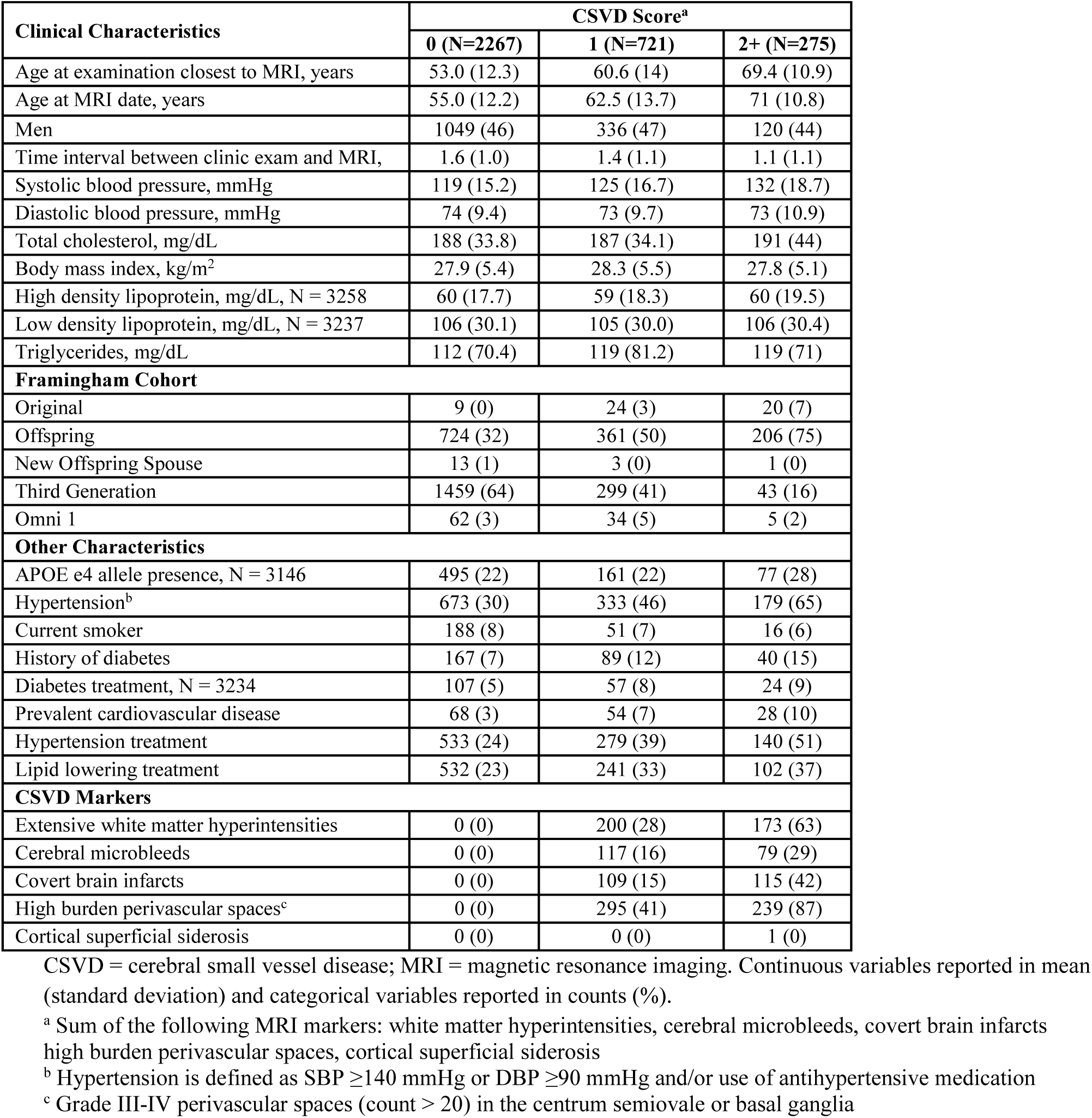
Sample characteristics stratified by CSVD score.

**Table 2.** Crude incidence rates for outcomes stratified by CSVD burden.

| Outcome | Measure (total N) | CSVD Score<sup>a</sup> |  |  |
| --- | --- | --- | --- | --- |
|  |  | 0 (N=2267) | 1 (N=721) | 2+ (N=275) |
| Composite Outcome<sup>b</sup> | N Events | 187 | 163 | 104 |
|  | Incidence rate (95% CI) | 9.78 (8.48, 11.29) | 27.71 (23.77, 32.31) | 49.86 (41.14, 60.43) |
| Stroke | N Events | 53 | 48 | 28 |
|  | Incidence rate (95% CI) | 2.75 (2.10, 3.60) | 7.99 (6.02, 10.60) | 13.1 (9.04, 18.97) |
| Myocardial Infarction | N Events | 39 | 24 | 13 |
|  | Incidence rate (95% CI) | 1.78 (1.30, 2.44) | 3.53 (2.36, 5.26) | 5.23 (3.03, 9.00) |
| Mortality | N Events | 124 | 124 | 89 |
|  | Incidence rate (95% CI) | 5.62 (4.71, 6.70) | 17.82 (14.95, 21.25) | 34.97 (28.41, 43.04) |

CI = confidence interval; CSVD = cerebral small vessel disease; MRI = magnetic resonance imaging. Incidence rates per 1,000 person-years.
<sup>a</sup> Sum of the following MRI markers: white matter hyperintensities, cerebral microbleeds, covert brain infarcts high burden perivascular spaces, cortical superficial siderosis.
<sup>b</sup> Includes all-cause mortality, stroke, and MI

**Table 3.** Cox regression results of associations with cerebral small vessel disease score.

| Outcome | Model | CSVD Score<sup>a</sup> | Hazard Ratio (95% CI) | p-value |
| --- | --- | --- | --- | --- |
| Composite Outcome<sup>b</sup> | 1 | 0 | ref | - |
|  |  | 1 | 1.48 (1.19, 1.85) | < 0.01 |
|  |  | 2+ | 1.52 (1.17, 1.97) | < 0.01 |
|  | 2 | 0 | ref | - |
|  |  | 1 | 1.46 (1.17, 1.81) | < 0.01 |
|  |  | 2+ | 1.48 (1.14, 1.93) | < 0.01 |
| Mortality | 1 | 0 | ref | - |
|  |  | 1 | 1.53 (1.18, 1.98) | < 0.01 |
|  |  | 2+ | 1.65 (1.24, 2.21) | < 0.01 |
|  | 2 | 0 | ref | - |
|  |  | 1 | 1.49 (1.15, 1.93) | < 0.01 |
|  |  | 2+ | 1.64 (1.23, 2.20) | < 0.01 |
| Stroke | 1 | 0 | ref | - |
|  |  | 1 | 1.59 (1.06, 2.40) | 0.02 |
|  |  | 2+ | 1.64 (1.01, 2.68) | 0.04 |
|  | 2 | 0 | ref | - |
|  |  | 1 | 1.56 (1.04, 2.35) | 0.03 |
|  |  | 2+ | 1.61 (0.99, 2.64) | 0.06 |

CI = confidence interval; CSVD = cerebral small vessel disease; MRI = magnetic resonance imaging.
Model 1 adjusts for age, sex, Framingham cohort, and time interval between clinic examination and MRI.
Model 2 additionally adjusts for vascular risk factors including hypertension, smoking status, diabetes, lipid lowering treatment, and body mass index.
<sup>a</sup> Sum of the following MRI markers: white matter hyperintensities, cerebral microbleeds, covert brain infarcts high burden perivascular spaces, cortical superficial siderosis.
<sup>b</sup> Includes all-cause mortality, stroke, and MI

The primary outcome was time to first event, defined as the earliest occurrence of death of any cause, MI, or stroke. If no event occurred over the course of follow-up, participants were censored at the time they were last known to be event-free. Time to each individual event were also evaluated separately as secondary outcomes. Participants could contribute to more than one individual outcome if they experienced multiple events. Incidence rates for all outcomes were calculated over the study period following MRI acquisition and measured by 1000 person years.

Multivariable Cox proportional hazard regression analyses were used to relate the multi-marker CSVD score to time to first event (and each individual event), estimating hazard ratios (HRs) and their 95% confidence intervals. CSVD score was treated as a categorical variable, with a score 0 (no CSVD markers) serving as the reference group. The proportional hazards assumption was assessed using weighted Schoenfeld residuals.

The primary model (model 1) adjusted for age, sex, FHS cohort, and time interval between clinic examination and MRI. The secondary model (model 2) additionally adjusted for vascular risk factors including hypertension, smoking status, diabetes, lipid lowering treatment, and BMI. In exploratory analyses, we assessed effect modification by sex and the association of each CSVD marker individually on the time to event outcomes. All statistical analyses were performed using SAS version 9.4 (SAS Institute, Cary, NC), and a two-sided p-value < 0.05 was considered statistically significant.

## Results

### Descriptive Statistics

Baseline characteristics stratified by exposure (CSVD score) are presented in Table 1. The majority of participants underwent 1.5T scans (N = 3,085; 90%), while 178 (10%) underwent 1.0T scans. Of the 3,263 participants in the sample (mean age 58.0 years [SD: 13.4], 54% women), 996 (31%) had CSVD scores greater than 0. Participants with higher CSVD scores were older, more likely to have diabetes and hypertension, and had greater systolic blood pressures and total cholesterol levels.

A comparison of baseline characteristics for included versus excluded participants is shown in Table 1 in supplemental material. The excluded participants tended to be older, had higher proportions of hypertension and CVD, and higher proportions of having been on diabetes, hypertension, and lipid lowering treatments. In addition, they had higher prevalence of WMH, CMB, CBI, PVS.

### Adverse Event Incidence

Over a median follow up of 8.4 years (IQR: 6.2 - 10.0), 454 participants (13.9%) experienced mortality, MI, or stroke. Mortality was the most common outcome (N = 337, 10%), followed by stroke (N = 129, 4%) and MI (N = 76, 2%). Crude incidence rates of the composite outcome, stroke, MI and mortality increased steadily as CSVD score increased (Table 2). These findings are reflected in Kaplan-Meier curves plotting survival probability as a function of follow-up time (Figure 2).

**Figure 2.**
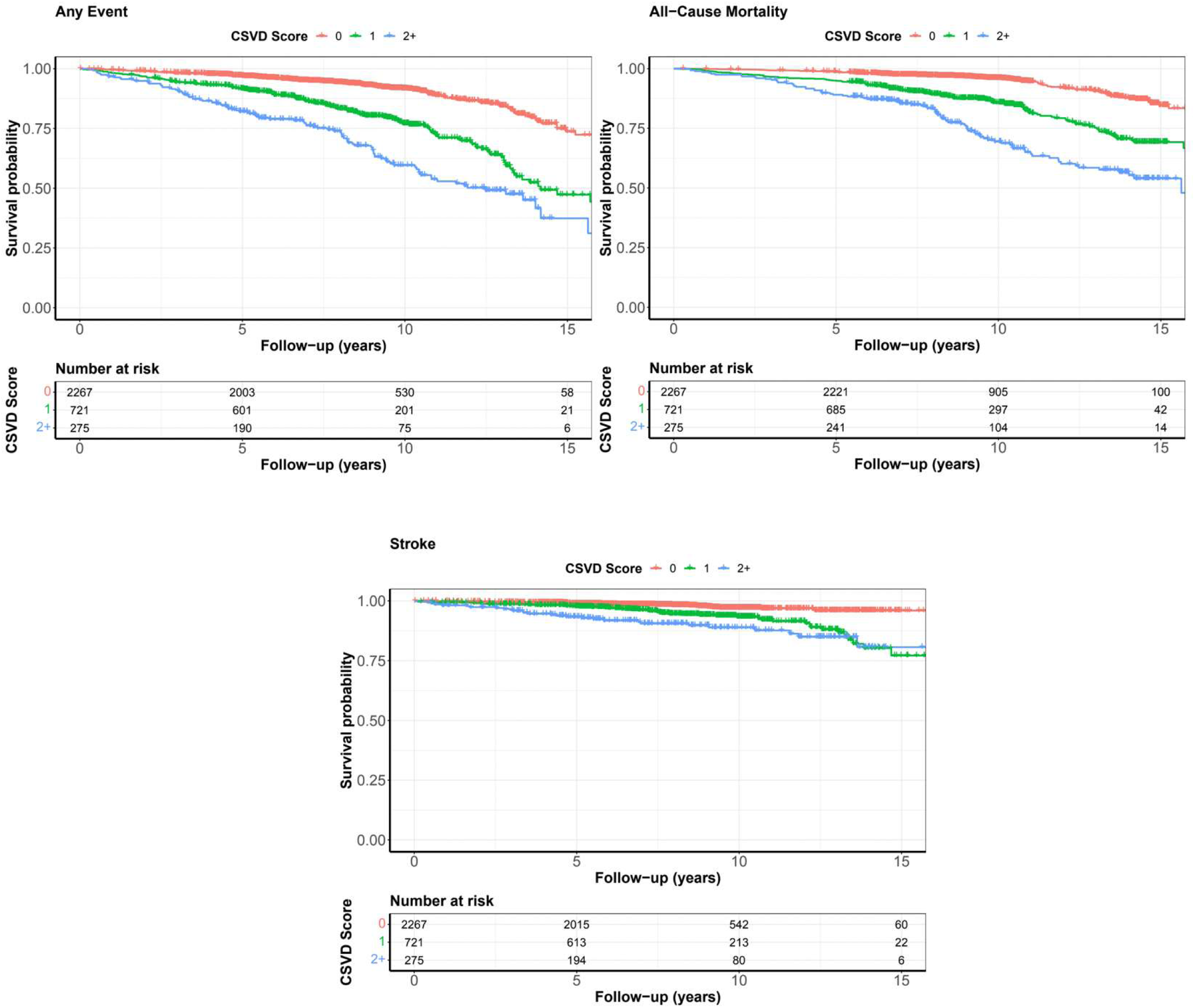
Kaplan-Meier curves for event incidence stratified by CSVD score. Any event is the composite outcome comprised of all-cause mortality, MI, and stroke.

### Multivariable Analyses

Multivariable analyses are reported in Table 3. The primary results reported below are based on model 1 (adjusted for age, sex, FHS cohort, and time interval between clinic examination and MRI). Further adjustments for vascular risk factors, including hypertension, smoking status, diabetes, lipid lowering treatment, and BMI (Model 2) did not change the associations.

We observed statistically significant associations for the composite outcome. Participants with a CSVD score of 1 had approximately a 48% increased risk of experiencing any event (HR 1.48, 95% CI 1.19 – 1.85), and those with a score of 2 had a 52% greater risk (HR 1.52, 95% CI 1.17 – 1.97) compared to those with CSVD score 0.

For the individual outcome of mortality, there was a significantly higher risk with a dose-effect relation, showing increasing risk as the CSVD score increased. A CSVD score of 1 was correlated with a 53% higher risk (HR 1.53, 95% CI 1.19 – 1.98), a score of 2 with a 65% higher risk (HR 1.65, 95% CI 1.24 – 2.21).

Our analyses showed a similar dose-effect relationship between CSVD burden and stroke (score 1: HR 1.59, 95% CI 1.06 – 2.40; score 2: HR 1.64, 95% CI 1.01 – 2.68).

For the individual outcome of MI, we were limited due to the small number of events in certain subgroups so results were not reported.

### Effect Modification by Sex

Results from Cox regression models stratified by sex are contained in Table 2 in the Supplemental Material. A CSVD score of 1 was associated with a similar and significant increase in risk of the composite event for both men (HR = 1.45, 95% CI: 1.08–1.96) and women (HR = 1.47, 95% CI: 1.06–2.04). For score 2+, there was also a similarly higher risk in both men (HR = 1.54, 95% CI: 1.07–2.20) and women (HR = 1.48, 95% CI: 1.01–2.17).

Among women, CSVD scores of 1 (HR = 1.64, 95% CI: 1.12–2.41) and 2+ (HR = 1.67, 95% CI: 1.08–2.58) were significantly associated with increased risk of all-cause mortality. In men, only score 2+ reached statistical significance (HR = 1.53, 95% CI: 1.03–2.28), while score 1 (HR = 1.38, 95% CI: 0.97–1.96) associated with a moderate but non-significant increase in risk. Numerically, the risk was slightly higher for women than men.

There were no significant associations with risk of stroke in either men or women.

### CSVD Individual Markers

The analyses between the adverse outcomes and each individual CSVD marker is shown in Table 3 in Supplemental Material. High burden of PVS and extensive WMH were significantly associated with increased risk of the composite outcome, with hazard ratios of 1.51 (95% CI: 1.23–1.86) and 1.46 (95% CI: 1.14–1.89), respectively. Both markers were also associated with increased risk of mortality, with HRs of 1.59 (95% CI: 1.26–2.00) for PVS and 1.67 (95% CI: 1.26–2.22) for WMH.

Cerebral microbleeds were the only marker significantly associated with increased risk of stroke (HR = 1.75; 95% CI: 1.06-2.88).

## Discussion

In our sample of healthy community-dwelling individuals, we found that a multi-marker CSVD score is associated with increased risk of any event, mortality, and stroke. We observed differences in associations between men and women and found that individual CSVD markers showed varying associations with mortality and stroke.

Our study offers novel insights into the relation of subclinical CSVD and major cardiovascular events in healthy, community dwelling individuals. Our results position subclinical CSVD as a potential intermediate biomarker that identifies individuals at increased risk for future events in an otherwise healthy population.

The link between clinical stroke and CSVD is well-established in the literature,^7,8^ which our study further supports. In terms of cardiovascular events, a few previous studies provide important context. A previous cross-sectional analysis of the Rotterdam study showed that unrecognized MI was related to higher prevalence of CSVD in men.^33^ ARIC study investigators found correlations between family history of coronary heart disease (CHD) and CSVD.^34^ Another study measured the association between ideal cardiovascular health (measured by LS7) and CSVD risk, finding that excellent CV health was associated with lower total CSVD burden.^35^ Notably, these studies looked at CSVD score as the outcome of interest, which is how it is traditionally looked at. Our study takes the reverse approach and analyzes CSVD as an exposure, offering perspectives on its role in predicting future adverse outcome risk.

Few studies have analyzed CSVD as an exposure in relation to other cardiovascular outcomes. In one study, cardiovascular and aortic pathologies were shown to be more prevalent in small vessel dementia than other vascular dementia cases.^36^ Analyzing CSVD in this light becomes important, especially in recent years. The increasing availability of brain MRI has resulted in increasing recognition of CSVD in subclinical stages, which—in view of its occurrence years to decades prior to clinical outcomes—offers a tremendous opportunity for prevention of vascular events.

Our study presents results assessing the relationship between subclinical CSVD and adverse cardiovascular events and mortality in a large cohort of asymptomatic, community-dwelling individuals. Only a few studies have analyzed this relationship, and those that have done so have focused on specific clinical contexts. One study found a strong association between at least one CSVD marker presence and major adverse cardiac and cerebrovascular events (MACCE)—a composite endpoint but with the addition of a variable for any revascularization procedure—only in patients with pre-existing hypertension.^15^ In contrast, our study used a composite CSVD score, capturing disease severity, as well as using a broader population of individuals. Another study examined high CSVD burden, also using a composite score, in patients with acute coronary syndrome and similarly found a strong association with MACCE.^37^ In patients with coronary artery dissections, a prior study found that CSVD burden did not relate to overall 6-month risk of major adverse cardiovascular events, but suggested it may be associated with later event occurrence.^38^

These studies all analyzed CSVD and cardiovascular outcomes exclusively in populations with pre-existing conditions. Our study, by our knowledge, presents a novel analysis of CSVD and adverse events in a larger sample of community-dwelling participants, showing how this relationship extends even to healthy individuals. In addition, our study expands current knowledge by including a larger sample size, broader age range, and a longer follow-up time. Additionally, the Framingham cohort tends to have a lower prevalence of traditional cardiovascular risk factors, further demonstrating the relevance of these findings even in individuals considered healthy.

This study also found some differences in risk between men and women. There was a significant and higher risk of mortality in women with the highest CSVD burden, while higher CSVD burden in men was significantly associated with increased stroke risk. Prior studies looking at sex-specific differences have reached varying conclusions, with some showing higher rates of major adverse cardiovascular events, stroke, and mortality in women while others show higher rates in men.^39–41^ Further research is needed to see if our findings are clinically relevant.

The mechanisms by which CSVD relates to these adverse events are not fully elucidated, but likely involve many shared pathophysiological aspects. Several common modifiable vascular risk factors are linked to CSVD and cardiovascular events; however, simple occurrence of vascular risk factors is not enough to explain the relation as we observed significant increases in risk despite adjustment for cardiovascular risk factors in our analyses. It is likely that a more complex relation exists which includes consideration of long-term achievement of risk factor control, medication and medication classes used. As such, burden of CSVD may reflect more accurately these exposures and treatment effects.

Small vessel disease is likely a systemic process affecting multiple organ systems. From a biological standpoint, mechanisms such as endothelial dysfunction and vascular inflammation are likely to contribute to CSVD and its relation to adverse cardiovascular events.^42–45^ These processes can impair vascular autoregulation^46^ and consequent organ perfusion as well as alter the balance of pro and antithrombotic systems, eventually leading to acute clinical events.

This study has limitations that warrant consideration. Despite the large sample size, there were a limited number of events for MI, which limited power for subgroup analysis with this outcome. In addition, this study had a predominantly White sample, which limits generalizability to some extent. The study was observational in design and hence we cannot exclude residual confounding and cannot establish causal relations. The participants excluded from this study had a higher vascular risk factor profile than those included. Among the strengths of this study are its prospective cohort design with long-term follow-up, the inclusion of a large sample, expanding the results to not only those with pre-existing clinical conditions but showing that the relation between CSVD and these outcomes exists even in the general population. Assessment of the CSVD markers on MRI was done blinded to all clinical and demographic characteristics and with high reproducibility of ratings.

## Conclusion

In this large prospective cohort study of community-dwelling individuals, higher burden of CVSD was related to higher risk of adverse events. The association showed a dose-response relation and was independent of vascular risk factors. These findings suggest that CSVD burden may be useful for risk stratification and prevention of adverse outcomes. Assessing CSVD burden in healthy, community-dwelling individuals could help identify those with an increased risk of adverse events, allowing for closer monitoring and earlier intervention to enhance preventive strategies for life-threatening outcomes.

## Data Availability

Requests to access data from the Framingham Heart Study can be made at https://www.framinghamheartstudy.org/fhs?for?researchers.

## Acknowledgements and Funding Sources

Sources of Funding:

This work (design and conduct of the study, collection and management of the data) was supported by the Framingham Heart Study’s National Heart, Lung, and Blood Institute contract (N01-HC-25195; HHSN268201500001I) and by grants from the National Institute of Neurological Disorders and Stroke (R01-NS017950-40, R21 NS135268), the National Institute on Aging (R01 AG059725; AG008122; AG054076; K23AG038444; R03 AG048180-01A1; AG033193); NIH grant (P30 AG010129).

This work was also supported by the Boston University Undergraduate Research Opportunities Program.

